# Concurrent human antibody and T_H_1 type T-cell responses elicited by a COVID-19 RNA vaccine

**DOI:** 10.1101/2020.07.17.20140533

**Authors:** Ugur Sahin, Alexander Muik, Evelyna Derhovanessian, Isabel Vogler, Lena M. Kranz, Mathias Vormehr, Alina Baum, Kristen Pascal, Jasmin Quandt, Daniel Maurus, Sebastian Brachtendorf, Verena Lörks, Julian Sikorski, Rolf Hilker, Dirk Becker, Ann-Kathrin Eller, Jan Grützner, Carsten Boesler, Corinna Rosenbaum, Marie-Cristine Kühnle, Ulrich Luxemburger, Alexandra Kemmer-Brück, David Langer, Martin Bexon, Stefanie Bolte, Katalin Karikó, Tania Palanche, Boris Fischer, Armin Schultz, Pei-Yong Shi, Camila Fontes-Garfias, John L. Perez, Kena A. Swanson, Jakob Loschko, Ingrid L. Scully, Mark Cutler, Warren Kalina, Christos A. Kyratsous, David Cooper, Philip R. Dormitzer, Kathrin U. Jansen, Özlem Türeci

**Affiliations:** BioNTech, An der Goldgrube 12, 55131 Mainz, Germany; TRON gGmbH – Translational Oncology at the University Medical Center of the Johannes Gutenberg, University Freiligrathstraße 12, 55131 Mainz, Germany; Pfizer, 401 N Middletown Rd., Pearl River, NY 10960, U.S.A.; Regeneron Pharmaceuticals, Inc., 777 Old Saw Mill River Rd, Tarrytown, NY 10591, U.S.A.; University of Texas Medical Branch, Galveston, TX 77555, U.S.A.; CRS Clinical Research Services Mannheim GmbH, Grenadierstrasse 1, 68167 Mannheim, Germany; Bexon Clinical Consulting LLC, Upper Montclair, NJ 07043, U.S.A.

## Abstract

An effective vaccine is needed to halt the spread of the SARS-CoV-2 pandemic. Recently, we reported safety, tolerability and antibody response data from an ongoing placebo-controlled, observer-blinded phase 1/2 COVID-19 vaccine trial with BNT162b1, a lipid nanoparticle (LNP) formulated nucleoside-modified messenger RNA encoding the receptor binding domain (RBD) of the SARS-CoV-2 spike protein. Here we present antibody and T cell responses after BNT162b1 vaccination from a second, non-randomized open-label phase 1/2 trial in healthy adults, 18-55 years of age. Two doses of 1 to 50 µg of BNT162b1 elicited robust CD4^+^ and CD8^+^ T cell responses and strong antibody responses, with RBD-binding IgG concentrations clearly above those in a COVID-19 convalescent human serum panel (HCS). Day 43 SARS-CoV-2 serum neutralising geometric mean titers were 0.7-fold (1 µg) to 3.5-fold (50 µg) those of HCS. Immune sera broadly neutralised pseudoviruses with diverse SARS-CoV-2 spike variants. Most participants had T_H_1 skewed T cell immune responses with RBD-specific CD8^+^ and CD4^+^ T cell expansion. Interferon (IFN)γ was produced by a high fraction of RBD-specific CD8^+^ and CD4^+^ T cells. The robust RBD-specific antibody, T-cell and favourable cytokine responses induced by the BNT162b1 mRNA vaccine suggest multiple beneficial mechanisms with potential to protect against COVID-19.

## Introduction

In December 2019, the novel coronavirus SARS-CoV-2 emerged in China causing coronavirus disease 2019 (COVID-19), a severe, acute respiratory syndrome with a complex, highly variable disease pathology. On 11 March 2020, the World Health Organization (WHO) declared the SARS-CoV-2 outbreak a pandemic. As of 29 June 2020, over 10 million cases have been reported worldwide, with deaths approaching half a million^1^.

The high and worldwide impact on human society calls for the rapid development of safe and effective therapeutics and vaccines^2^.

Messenger RNA (mRNA) vaccine technology allows to deliver precise genetic information encoding a viral antigen together with intrinsic adjuvant effect to antigen presenting cells^3^. The prophylactic effectiveness of this technology has been proven in preclinical models against multiple viral targets^4–6^. LNP- and liposome-formulated RNA vaccines for prevention of infectious diseases and for treatment of cancer have been shown in clinical trials to be safe and well-tolerated^7^. mRNA is transiently expressed and does not integrate into the genome. It is molecularly well defined, free of animal-origin materials and synthesized by an efficient, cell-free *in vitro* transcription process from DNA templates^4,8,9^. The fast and highly scalable mRNA manufacturing and lipid-nanoparticle (LNP) formulation processes enable rapid production of many vaccine doses^5,6,10^, making it suitable for rapid vaccine development and pandemic vaccine supply.

Two Phase 1/2 umbrella trials in Germany and the US investigate several LNP-encapsulated RNA vaccine candidates developed in ‘Project Lightspeed’. Recently, we have reported interim data obtained in the US trial (NCT04368728) for the most advanced candidate BNT162b1^11^. BNT162b1 encodes the receptor-binding domain (RBD) of the SARS-CoV-2 spike protein, a key target of neutralising antibodies. The RBD antigen expressed by BNT162b1 is fused to a T4 fibritin-derived “foldon” trimerisation domain to increase its immunogenicity by multivalent display^12^. The RNA is optimized for high stability and translation efficiency^13,14^ and incorporates 1-methyl-pseudouridine instead of uridine to dampen innate immune sensing and to increase mRNA translation *in vivo*^15^. In the placebo-controlled, observer-blinded US trial, dosages of 10 µg, 30 µg (prime and boost doses 21 days apart for both dose levels) and 100 µg (prime only) were administered. No serious adverse events were reported. Local injection site reactions and systemic events (mostly flu-like symptoms) were dose-dependent, generally mild to moderate, and transient. RBD-binding IgG concentrations and SARS-CoV-2 neutralising titers in sera increased with dose level and after a second dose. Fourteen days after the boost, geometric mean neutralising titers reached 1.9-to 4.6-fold of a panel of COVID-19 convalescent human sera.

This study now complements our previous report with available data from the German trial (NCT04380701, EudraCT: 2020-001038-36), providing a detailed characterisation of antibody and T-cell immune responses elicited by BNT162b1 vaccination.

## Results

### Study design and analysis set

Between 23 April 2020 and 22 May 2020, 60 participants were vaccinated with BNT162b1 in Germany. Twelve participants per 1 µg, 10 μg, 30 μg, and 50 µg dose level groups received a first dose on Day 1 and were boosted on Day 22 (except for one individual each in the 10 and 50 µg dose-level cohort who discontinued due to non-study drug related reasons), and 12 participants received a 60 μg prime dose on Day 1 only (Extended Data Figure 1). The study population consisted of healthy males and non-pregnant females with a mean age of 41 years (range 18 to 55 years) with equal gender distribution. Most participants were Caucasian (96.7%) with one African American and one Asian participant (1.7% each). Preliminary data analysis focused on immunogenicity (Extended Data Table 1).

### Preliminary available safety and tolerability data

Briefly, no serious adverse events (SAE) and no withdrawals due to related adverse events (AEs) were observed for any dose. Similar to the U.S. trial, most reported solicited events in the 10 µg and 30 µg groups were reactogenicity (*e*.*g*., fatigue and headache), with a typical onset within the first 24 hours after immunisation. Injection site reactions within 7 days of the prime or boost were mainly pain and tenderness (Extended Data Figure 2). Symptomatology was mostly mild or moderate with occasional severe (Grade 3) reactogenicity such as fever, chills, headache, muscle pain, joint pain, injection site pain, and tenderness within 7 days after each dose. Reactogenicity resolved spontaneously, or could be managed with simple measures (*e*.*g*. paracetamol). Based on the reactogenicity reported after the 50 µg boost dose, a second 60 μg dose was not administered to participants who had received an initial 60 μg dose.

Whereas no relevant change in routine clinical laboratory values occurred after BNT162b1 vaccination, a transient increase in C-reactive protein (CRP) and temporary reduction of blood lymphocyte counts were observed in a dose-dependent manner in vaccinated participants (Extended Data Figure 3). CRP is a well-known inflammatory serum protein previously described as biomarker for various infectious disease vaccines and an indicator of vaccine adjuvant activity^16–19^. Based on our previous clinical experience with RNA vaccines the transient decrease in lymphocytes is likely attributable to innate immune stimulation-related redistribution of lymphocytes into lymphoid tissues^20^. Both parameters are considered pharmacodynamics markers for the mode-of-action of RNA vaccines.

### Vaccine-induced antibody response

RBD-binding IgG concentrations and SARS-CoV-2 neutralising titers were assessed at baseline, 7 and 21 days after the BNT162b1 priming dose (Days 8 and 22), and 7 and 21 days after the boosting dose (Days 29 and 43), except for the 60 µg cohort, which received a priming dose only (Figure 1).

**Figure 1.**
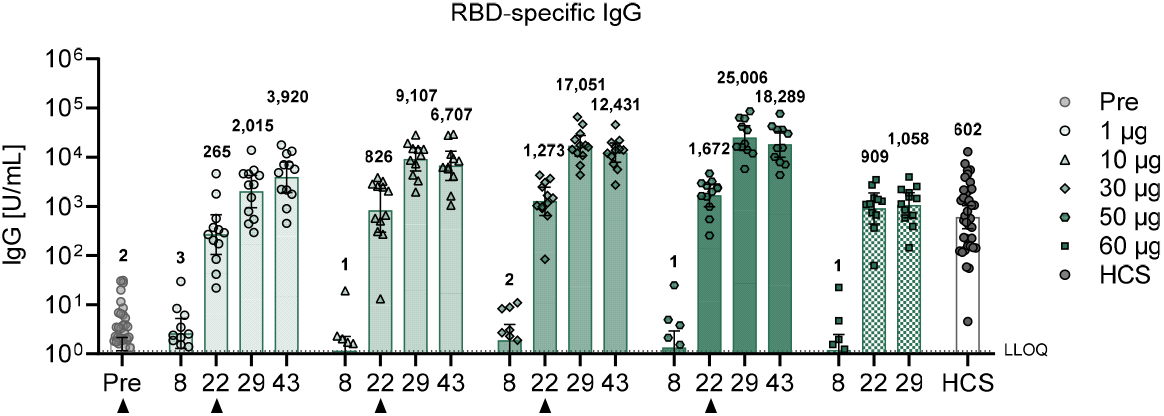
BNT162b1-induced IgG concentrations. Participants were immunised with BNT162b1 on Days 1 (all dose levels) and 22 (all dose levels except 60 µg) (*n*=12 per group, from Day 22 on *n*=11 for the 10 µg and 50 µg cohort). Sera were obtained on Day 1 (Pre prime) and on day 8, 22 (pre boost), 29 and 43. Pre-dose responses across all dose levels were combined. Human COVID-19 convalescent sera (HCS, *n*=38) were obtained at least 14 days after PCR-confirmed diagnosis and at a time when the donors were no longer symptomatic. For RBD-binding IgG concentrations below the lower limit of quantification (LLOQ = 1.15), LLOQ/2 values were plotted. Arrowheads indicate vaccination. Chequered bars indicate that no boost immunisation was performed. Values above bars are geometric means with 95% confidence intervals. At the time of submission, Day 43 data were pending for all participants of the 60 µg cohort.

Immunised participants showed a strong, dose-dependent vaccine-induced antibody response. Twenty-one days after the priming dose (for the four dose levels ranging from 1-50 µg), geometric mean concentrations (GMCs) of RBD-binding IgG had increased in a dose level dependent manner, with GMCs ranging from 265-1,672 U/mL (Figure 1). Seven days after the boosting dose (Day 29), RBD-binding IgG GMCs in participants vaccinated with 1-50 µg BNT162b1 showed a strong, dose-level dependent booster response ranging from 2,015-25,006 U/mL. At Day 43 (21 days after boost), RBD-binding antibody GMCs were in the range of 3,920-18,289 U/mL in BNT162b1 vaccinated individuals as compared to a GMC of 602 U/mL measured in a panel of convalescent sera from 38 SARS-CoV-2 infection patients. The patients were 18-83 years of age, and sera were drawn at least 14 days after PCR-confirmed diagnosis. In the 60 µg dose-level cohort, which received a priming dose only, RBD-binding IgG GMCs were 1,058 U/mL by Day 29, indicating that a boosting dose to increase antibody titers may be necessary.

SARS-CoV-2 neutralising antibody geometric mean titers (GMTs) increased modestly in a dose-dependent manner 21 days after the priming dose (Figure 2a). Substantially higher serum-neutralising GMTs were achieved 7 days after the booster dose, reaching 36 (1 µg dose level), 158 (10 µg dose level), 308 (30 µg dose level), and 578 (50 µg dose level), compared to 94 for the convalescent serum panel. On Day 43 (21 days after the boost), the neutralising GMTs decreased (with exception of the 1 µg dose level). Neutralising antibody GMTs were strongly correlated with RBD-binding IgG GMC (Figure 2b). In summary, antibody responses elicited by BNT162b1 in study BNT162-01 largely mirrored those observed in the U.S. study^11^.

**Figure 2.**
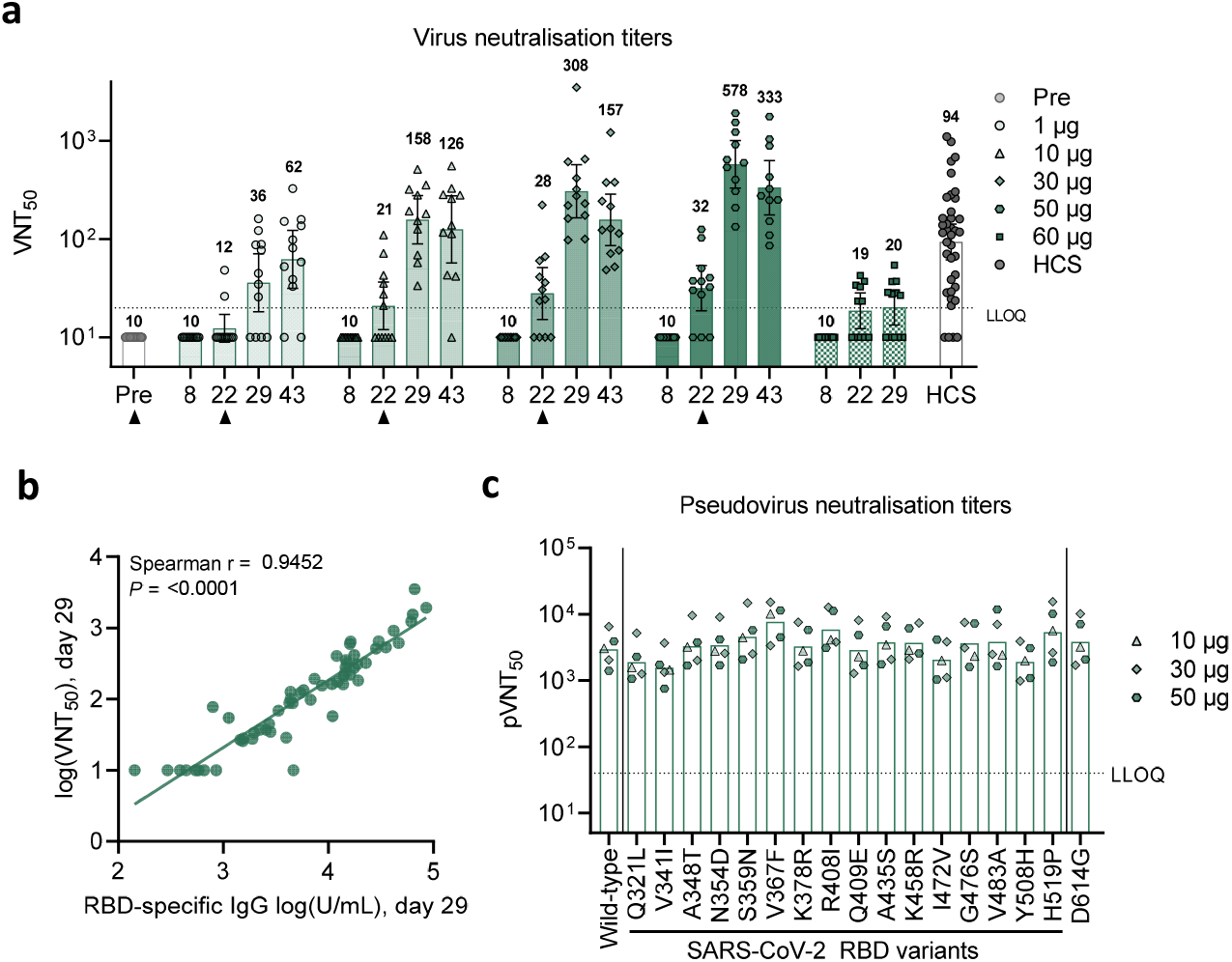
BNT162b1-induced virus neutralisation titers. The vaccination schedule and serum sampling are described in Figure 1. **a**, SARS-CoV-2 50% neutralisation titers (VNT_50_) in immunised participants and COVID-19 convalescent patients (HCS). For values below the lower limit of quantification (LLOQ) = 20, LLOQ/2 values were plotted. Arrowheads indicate days of immunisation. Chequered bars indicate that no boost immunisation was performed. Geometric mean titers (values above bars) with 95% confidence interval are shown. At the time of submission, Day 43 data were pending for all participants of the 60 µg cohort, **b**, Correlation of RBD-binding IgG geometric mean concentrations (GMC) (as in Figure 1) with VNT_50_ on Day 29 (all evaluable participant sera). Nonparametric Spearman correlation. **c**, Pseudovirus 50% neutralisation titers (pVNT_50_) across a pseudovirus panel displaying 17 SARS-CoV-2 spike protein variants including 16 RBD mutants and the dominant spike protein variant D614G (dose levels 10, 30 and 50 µg, *n*=1-2 each; Day 29). Lower limit of quantification (LLOQ) = 40. Geometric mean titers are displayed.

To demonstrate the breadth of the neutralising response, a panel of 16 SARS-CoV-2 RBD variants identified through publicly available information^21^ and the dominant (non-RBD) spike variant D614G^22^ was evaluated in pseudovirion neutralisation assays. Sera collected 7 days after the second dose of BNT162b1 showed high neutralising titers to each of the SARS-CoV-2 spike variants (Figure 2c).

### Vaccine-induced T cell responses

CD4^+^ and CD8^+^ T cell responses in BNT162b1 immunised participants were characterised prior to priming vaccination (Day 1) and 7 days after boost vaccination (on Day 29) using direct *ex vivo* IFNγ ELISpot with peripheral blood mononuclear cells (PBMCs) from 36 participants across the 1 µg to 50 µg dose-level cohorts (Figure 3). In this assay, CD4^+^ or CD8^+^ T cell effectors were stimulated overnight with overlapping peptides representing the full-length sequence of the vaccine-encoded RBD. Of 36 participants, 34 (94.4%, including all participants treated with ≥ 10 µg BNT162b1) mounted RBD-specific CD4^+^ T cell responses. While the magnitude varied between individuals, participants with the strongest CD4^+^ T cell responses to RBD had more than 10-fold of the memory responses observed in the same participants when stimulated with cytomegalovirus (CMV), Epstein Barr virus (EBV), influenza virus and tetanus toxoid-derived immuno-dominant peptide panels (Figure 3a-c). No CD4^+^ T cell responses were detectable at baseline, except for one participant with a low number of preexisting RBD-reactive CD4^+^ T cells, which increased significantly after vaccination (normalised mean spot count from 63 to 1,519, in the 50 µg dose cohort). The strength of RBD-specific CD4^+^ T cell responses correlated positively with both RBD-binding IgG and with SARS-CoV-2 neutralising antibody titers (Extended Data Figure 4a, d), in line with the concept of intramolecular help^23^. The two participants lacking CD4^+^ response had no detectable virus neutralising titers (VNT_50_) (Extended Data Figure 4d).

**Figure 3.**
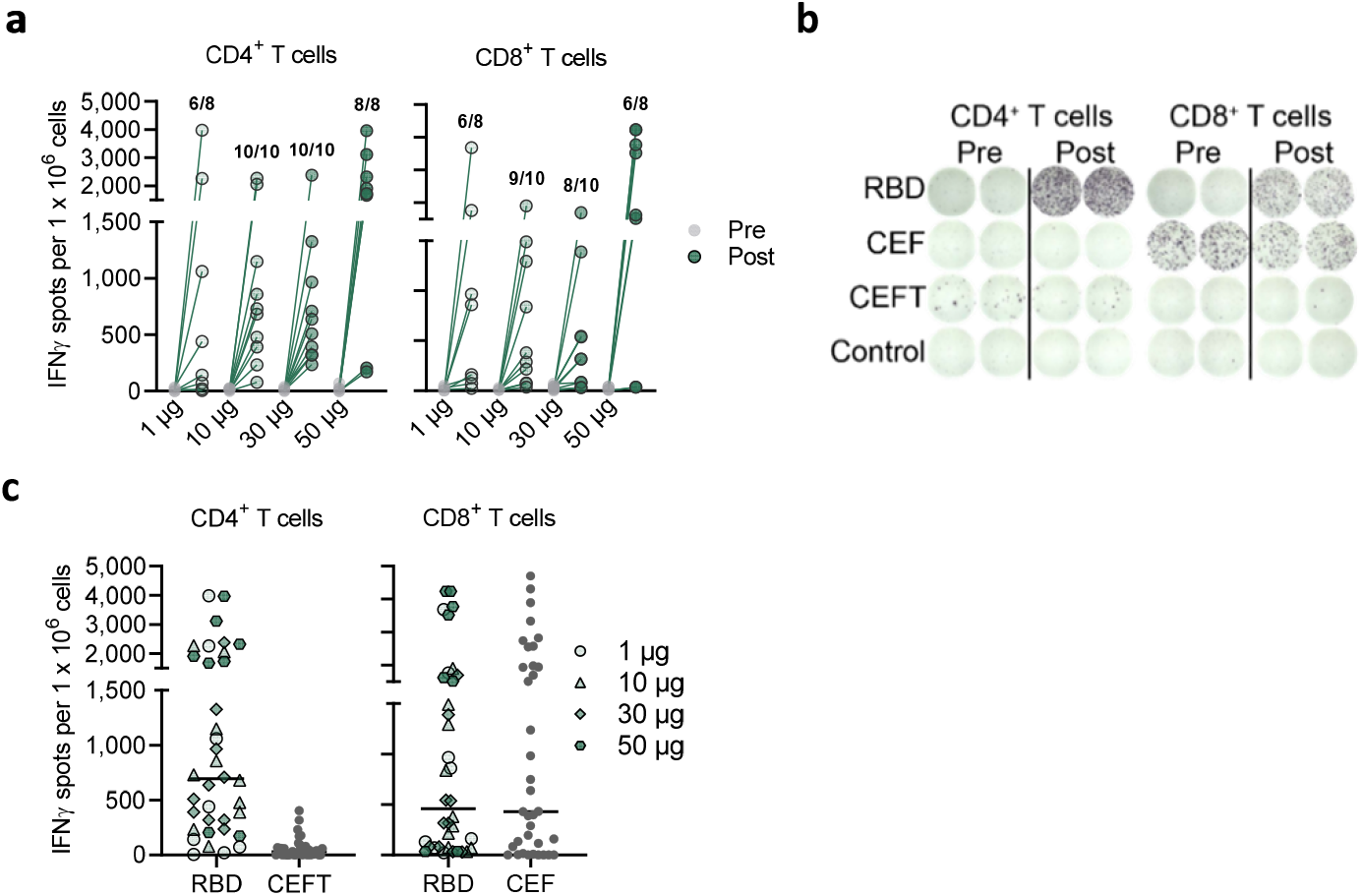
Frequency and magnitude of BNT162b1-induced CD4^+^ and CD8^+^ T-cell responses. The vaccination schedule is described in Figure 1. PBMCs obtained on Day 1 (Prevaccination) and on Day 29 (Postvaccination, 7 days after boost) (1 and 50 µg, *n*=8 each; 10 and 30 µg, *n*=10 each) were enriched for CD4^+^ or CD8^+^ T cell effectors and separately stimulated over night with an overlapping peptide pool representing the vaccine-encoded RBD for assessment in direct *ex vivo* IFNγ ELISpot. Common pathogen T-cell epitope pools CEF (CMV, EBV, influenza virus HLA class I epitopes) and CEFT (CMV, EBV, influenza virus, tetanus toxoid HLA class II epitopes) served to assess general T-cell reactivity, cell culture medium served as negative control. Each dot represents the normalized mean spot count from duplicate wells for one study participant, after subtraction of the medium-only control. **a**, Ratios above post-vaccination data points are the number of participants with detectable CD4^+^ or CD8^+^ T cell response within the total number of tested participants per dose cohort. **b**, Exemplary CD4^+^ and CD8^+^ ELISpot of a 10-µg cohort subject. **c**, RBD-specific CD4^+^ and CD8^+^ T cell responses in all prime/boost vaccinated participants and their baseline CEFT- and CEF-specific T-cell responses. Nonparametric Spearman correlation.

Among vaccine-induced CD8^+^ T cell responses (29/36 participants, 80.6%), strong responses were mounted by the majority of participants (Figure 3a) and were quite comparable with memory responses against CMV, EBV, influenza virus and tetanus toxoid in the same participants (Figure 3b, c). The strength of RBD-specific CD8^+^ T cell responses correlated positively with vaccine-induced CD4^+^ T cell responses, but did not significantly correlate with SARS-CoV-2 neutralising antibody titers (Extended Data Figure 4b, c).

Of note, although at 1 µg BNT162b1 the immunogenicity rate was lower (6/8 responding participants), the magnitude of vaccine-induced CD4^+^ and CD8^+^ T cells in some participants was almost as high as with 50 µg BNT162b1 (Figure 3a). To assess functionality and polarisation of RBD-specific T cells, cytokines secreted in response to stimulation with overlapping peptides representing the full length sequence of the vaccine encoded RBD were determined by intracellular staining (ICS) for IFNγ, IL-2 and IL-4 specific responses in pre- and post-vaccination PBMCs of 18 BNT162b1 immunised participants. RBD-specific CD4^+^ T cells secreted IFNγ, IL-2, or both, but did not secrete IL-4 (Figure 4 a-c). Similarly, fractions of RBD-specific CD8^+^ T cells secreted IFNγ^+^ and IL-2.

**Figure 4.**
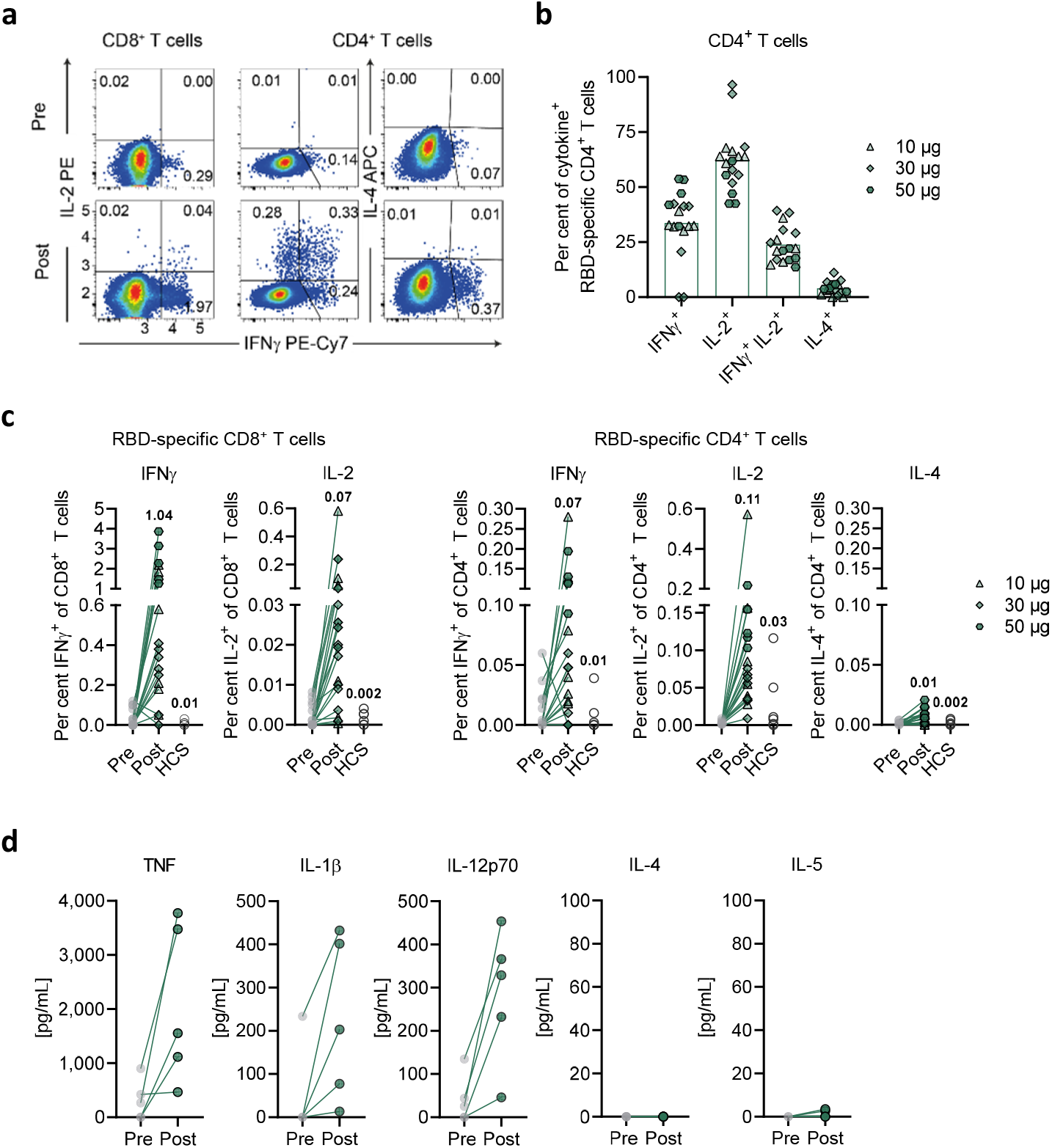
Cytokine polarisation of BNT162b1-induced T cells. The vaccination schedule and PBMC sampling are described in Figure 3. PBMCs of vaccinees and COVID-19 recovered donors (HCS *n*=6; in (c)) were stimulated over night with an overlapping peptide pool representing the vaccine-encoded RBD and analysed by flow cytometry (a-c) and bead-based immunoassay (d). **a**, Exemplary pseudocolor flow cytometry plots of cytokine-producing CD4^+^ and CD8^+^ T cells from a 10-µg cohort participant. **b**, RBD-specific CD4^+^ T cells producing the indicated cytokine as a fraction of total cytokine-producing RBD-specific CD4^+^ T cells. **c**, RBD-specific CD8^+^ (left) or CD4^+^ (right) T cells producing the indicated cytokine as a fraction of total circulating T cells of the same subset. One CD4 non-responder (<0.02%total cytokine producing T cells) from the 30-µg cohort was excluded in (b). Values above data points are the mean fractions across all dose cohorts. **d**, PBMCs from the 50-µg cohort. Each dot represents the mean from duplicate wells subtracted by the DMSO control for one study participant. Lower limits of quantification (LLOQ) were 6.3 pg/mL for TNF, 2.5 pg/mL for IL-1β, 7.6 pg/mL for IL-12p70, 11.4 pg/mL for IL-4 and 5.3 pg/mL for IL-5. Mean (b).

The mean fraction of RBD-specific T cells within total circulating T cells obtained by BNT162b1 vaccination was substantially higher than that observed in six participants who recovered from COVID-19. Fractions of RBD-specific IFNγ^+^ CD8^+^ T cells reached up to several percent of total peripheral blood CD8^+^ T cells (Figure 4c). Analysis of supernatants of PBMCs stimulated *ex vivo* with overlapping RBD peptides from a subgroup of five vaccinated participants detected proinflammatory cytokines TNF, IL-1β and IL-12p70, but neither IL-4 nor IL-5 (Figure 4d).

In summary, these findings indicate that BNT162b1 induces functional and proinflammatory CD4^+^/CD8^+^ T cell responses in almost all participants, with T_H_1 polarisation of the helper response.

## Discussion

We observed concurrent production of neutralising antibodies, activation of virus-specific CD4^+^ and CD8^+^ T cells, and robust release of immune-modulatory cytokines such as IFNγ, which represents a coordinated immune response to counter a viral intrusion (for review ^24^). IFNγ is a key cytokine for several antiviral responses. It acts in synergy with type I interferons to inhibit replication of SARS-CoV-2^25^. Patients with IFNγ gene polymorphism related to impaired IFNγ activity have been shown to display 5-fold increased susceptibility to SARS^26^. The robust production of IFNγ from CD8^+^ T cells indicates a favourable immune response with both anti-viral and immune-augmenting properties.

The detection of IFNγ, IL-2 and IL-12p70 but not IL-4 indicates a favorable T_H_1 profile and the absence of a potentially deleterious T_H_2 immune response. CD4^+^ and CD8^+^ T cells may confer long-lasting immunity against corona viruses as indicated in SARS-CoV-1 survivors, where CD8^+^ T-cell immunity persisted for 6-11 years^24,27^.

Some cases of asymptomatic virus exposure have been associated with cellular immune response without seroconversion indicating that SARS-CoV-2 specific T cells could be relevant in disease control even in the absence of neutralising antibodies^28^. Almost all vaccinated volunteers mounted RBD-specific T cell responses detected with an *ex vivo* ELISpot assay, which was performed without prior expansion of T cells that captures only high-magnitude T cell responses. Although the strength of the T-cell responses varied considerably between participants, we observed no clear dose dependency of the T-cell response strength using vaccine dose levels of 1 µg to 50 µg, indicating that stimulation and robust expansion of T cells might be accomplished at the lowest mRNA-encoded immunogen levels.

The study confirms the dose-dependency of RBD-binding IgG and neutralisation responses and reproduces our previous findings for the 10 and 30 µg dose levels of BNT162b1 in the US trial.

A notable observation is that two injections of BNT162b1 at a dose level as low as 1 µg are capable of inducing RBD-binding IgG levels higher than those observed in convalescent sera, and serum neutralising antibody titers that were still increasing up to Day 43. Considering that it is not known which neutralising antibody titer would be protective, and given the substantial T-cell responses we observed for some participants in the 1 µg cohort, a considerable fraction of individuals may benefit even from this lowest tested dose level.

A purely RBD-directed immunity might be considered prone to escape of the virus by single amino acid changes in this small domain. To address this concern, neutralisation assays were conducted with 17 pseudo-typed viruses, 16 of which enter cells using a spike with a different RBD variant found in circulating strains and one of which uses the dominant spike variant D614G. All 17 variants were efficiently neutralised by BNT162b1 immune sera.

Limitations of our clinical study include the small sample size and its restriction to participants below 55 years of age. Another constraint is that we did not perform further T cell analysis *e*.*g*. deconvolution of epitope diversity, characterisation of HLA restriction and TCR repertoire analysis before and after vaccination, due to the limited blood volumes that were available for biomarker analyses. Further, as vaccine-induced immunity can wane over time, it is important to study persistence of potentially protective immune responses over time. However, samples to assess persistence are not yet available but are planned per study protocol and will be reported elsewhere.

## Data Availability

The data that support the findings of this study are available from the corresponding author upon reasonable request. Upon completion of this clinical trial, summary-level results will be made public and shared in line with data sharing guidelines.

## Acknowledgements

We thank Mikael Dolsten, Pfizer Chief Scientific Officer, for advice during drafting of the manuscript. We thank C. Anders, C. Anft, N. Beckmann, K. Bissinger, G. Boros, P. Cienskowski, K. Clarke, C. Ecker, A. Engelmann, Y. Feuchter, L. Heesen, M. Hossainzadeh, S. Jägle, L. Jeck, O. Kahl, M. Knezovic, T. Kotur, M. Kretschmer, O. Pfante, J. Reinholz, L.-M. Schmid, R. Schulz, B. Stock, C. Müller, S. Murphy, G. Szabó and M. Vehreschild for technical support, project management and advice, and A. Finlayson and M. Rao for providing editorial assistance. We thank P. Koch and F. Groher for data management and analysis. We thank S. Liebscher and O. Kistner for expert advice. We thank Judith Absalon for manuscript advice. We thank the CRS Team (Mannheim and Berlin) for study conduct: S. Baumann, M. Berse, M. Casjens, B. Ehrlich, F. Seitz. We thank the Pfizer Vaccines Clinical Assays Team and the Pfizer Aviation Team for technical and logistical support of serology analyses.

## Author Contributions

U.S. conceived and conceptualised the work and strategy supported by Ö.T. Experiments were planned or supervised by E.D., C. F.-G, C.A.K., L.M.K., U.L., A.M., J.Q., P.-Y.S., and I.V.. A.B., D.C, M.C., C. F.-G, W.K., K.P., J.Q., I.S. and P.-Y.S. performed experiments. D.B., S.B., E.D., P.R.D., J.G., K.U.J., A.-K.E., L.M.K., M.-C.K., V.L., A.M., J.Q., J.S., I.V. and M.V. analysed data. D.M. planned and supervised dashboards for analysis of clinical trial data. R.H. was responsible for data normalization and adaption. C.B. and C.R. were responsible for biomarker and R&D program management. K.K. optimized the mRNA. M.B., S.B., B.F., A.K-B., D.L. and T.P., A.S., coordinated operational conduct of the clinical trial. J.L.P. advised on the trial, and J.L. and K.A.S advised on experiments. U.S., Ö.T., supported by M.B., E.D., P.R.D., K.U.J., L.M.K., A.M., I.V. and M.V., interpreted data and wrote the manuscript. All authors supported the review of the manuscript.

## Additional Information

Supplementary Information is available for this paper.

Correspondence and requests for materials should be addressed to Ugur Sahin.

## Materials and Methods

### Clinical trial design

Study BNT162-01 (NCT04380701) is an ongoing, first-in-human, Phase 1/2, open-label dose-ranging clinical trial to assess the safety, tolerability, and immunogenicity of ascending dose levels of various intramuscularly administered BNT162 mRNA vaccine candidates in healthy men and non-pregnant women 18 to 55 years (amended to add 56 -85 of age) of age. Key exclusion criteria included previous clinical or microbiological diagnosis of COVID-19; receipt of medications to prevent COVID-19; previous vaccination with any coronavirus vaccine; a positive serological test for SARS-CoV-2 IgM and/or IgG; and a SARS-CoV-2 NAAT-positive nasal swab; those with increased risk for severe COVID-19; and immunocompromised individuals. The primary endpoints of the study are safety and immunogenicity.

In the part of the study reported here, five dose levels (1 μg, 10 μg, 30 μg, 50 μg or 60 μg) of the BNT162b1 candidate were assessed at one site in Germany with 12 healthy participants per dose level in a dose-escalation/de-escalation design. Sentinel dosing was performed in each dose-escalation cohort. Progression in that cohort and dose escalation required data review by a safety review committee. Participants received a BNT162b1 prime dose on Day 1, and a boost dose on Day 22±2. Serum for antibody assays was obtained on Day 1 (pre-prime), 8±1 (post-prime), 22±2 (pre-boost), 29±3 and 43±4 (post-boost). PBMCs for T cell studies were obtained on Day 1 (pre-prime) and 29±3 (post-boost). Tolerability was assessed by patient diary. One subject of the 10 µg, and one subject of the 50 µg dose cohort left the study prior to the boosting immunisation due to withdrawal of consent and private reasons.

The presented data comprise the BNT162b1-immunised cohorts only and are based on a preliminary analysis with a data extraction date of 13 July 2020, focused on analysis of vaccine-induced immunogenicity (secondary endpoint) descriptively summarised at the various time points and on reactogenicity. All participants with data available were included in the immunogenicity analyses.

The trial was carried out in Germany in accordance with the Declaration of Helsinki and Good Clinical Practice Guidelines and with approval by an independent ethics committee (Ethik-Kommission of the Landesärztekammer Baden-Württemberg, Stuttgart, Germany) and the competent regulatory authority (Paul-Ehrlich Institute, Langen, Germany). All participants provided written informed consent.

### Manufacturing of RNA

BNT162b1 incorporates a Good Manufacturing Practice (GMP)-grade mRNA drug substance that encodes the trimerized SARS-CoV-2 spike glycoprotein RBD antigen. The RNA is generated from a DNA template by *in vitro* transcription in the presence of 1-methylpseudouridine-5’-triphosphate (m1YTP; Thermo Fisher Scientific) instead of uridine-5’-triphosphate (UTP). Capping is performed co-transcriptionally using a trinucleotide cap 1 analogue ((m_2_^7,3’-O^)Gppp(m^2’-O^)ApG; TriLink). The antigen-encoding RNA contains sequence elements that increase RNA stability and translation efficiency in human dendritic cells^13,14^. The mRNA is formulated with lipids to obtain the RNA-LNP drug product. The vaccine was transported and supplied as a buffered-liquid solution for IM injection and was stored at -80 °C.

### Proteins and peptides

A pool of 15-mer peptides overlapping by 11 amino acids and covering the whole sequence of the BNT162b1-encoded SARS-CoV-2 RBD, was used for *ex vivo* stimulation of PBMCs for flow cytometry, IFNγ ELISpot and cytokine profiling. CEF (CMV, EBV, influenza virus; HLA class I epitope peptide pool) and CEFT (CMV, EBV, influenza virus, tetanus toxoid; HLA class II epitope peptide pool) (both JPT Peptide Technologies) were used as controls for general T-cell reactivity.

### Human convalescent sera and PBMC panel

Human SARS-CoV-2 infection/COVID-19 convalescent sera (*n*=38) were drawn from donors 18-83 years of age at least 14 days after PCR-confirmed diagnosis and at a time when the participants were asymptomatic. Serum donors had symptomatic infections (*n*=35), or had been hospitalized (*n*=1). Sera were obtained from Sanguine Biosciences (Sherman Oaks, CA), the MT Group (Van Nuys, CA) and Pfizer Occupational Health and Wellness (Pearl River, NY). Human SARS-CoV-2 infection/COVID-19 convalescent PBMC samples (*n*=6) were collected from donors 41-79 years of age 45-59 days after PCR-confirmed diagnosis when donors were asymptomatic. PBMC donors had asymptomatic/mild infections (*n*= 4; clinical score 1 and 2) or had been hospitalized (*n*=2; clinical score 4 and 5). Blood samples were obtained from the Frankfurt University Hospital (Germany).

### Cell culture and primary cell isolation

Vero cells (ATCC CCL-81) and Vero E6 cells (ATCC CRL-1586) were cultured in Dulbecco’s modified Eagle’s medium (DMEM) with GlutaMAX(tm) (Gibco) supplemented with 10% fetal bovine serum (FBS) (Sigma-Aldrich). Cell lines were tested for mycoplasma contamination after receipt and before expansion and cryopreservation. Peripheral blood mononuclear cells (PBMCs) were isolated by Ficoll-Hypaque (Amersham Biosciences) density gradient centrifugation and cryopreserved prior to subsequent analysis.

### RBD binding IgG antibody assay

A recombinant SARS-CoV-2 RBD containing a C-terminal Avitag(tm) (Acro Biosystems) was bound to streptavidin-coated Luminex microspheres. Heat-inactivated participant sera were diluted 1:500, 1:5,000, and 1:50,000. Following an overnight incubation at 2-8 °C while shaking, plates were washed in a solution containing 0.05% Tween-20. A secondary fluorescently labelled goat anti-human polyclonal antibody (Jackson Labs) was added for 90 minutes at room temperature while shaking, before plates were washed once more in a solution containing 0.05% Tween-20. Data were captured as median fluorescent intensities (MFIs) using a Bioplex200 system (Bio-Rad) and converted to U/mL antibody concentrations using a reference standard curve (reference standard composed of a pool of five convalescent serum samples obtained >14 days post-COVID-19 PCR diagnosis and diluted sequentially in antibody-depleted human serum) with arbitrary assigned concentrations of 100 U/mL and accounting for the serum dilution factor. Three dilutions are used to increase the likelihood that at least one result for any sample will fall within the useable range of the standard curve. Assay results were reported in U/mL of IgG. The final assay results are expressed as the geometric mean concentration of all sample dilutions that produced a valid assay result within the assay range.

### SARS-CoV-2 neutralisation assay

The neutralisation assay used a previously described strain of SARS-CoV-2 (USA_WA1/2020) that had been rescued by reverse genetics and engineered by the insertion of an mNeonGreen (mNG) gene into open reading frame 7 of the viral genome^29^. This reporter virus generates similar plaque morphologies and indistinguishable growth curves from wild-type virus. Viral master stocks (2 × 10^7^ PFU/mL) were grown in Vero E6 cells as previously described^29^. With patient convalescent sera, the fluorescent neutralisation assay produced comparable results to the conventional plaque reduction neutralisation assay^30^. Serial dilutions of heat-inactivated sera were incubated with the reporter virus (2 × 10^4^ PFU per well to yield approximately a 10-30% infection rate of the Vero CCL81 monolayer) for 1 hour at 37 °C before inoculating Vero CCL81 cell monolayers (targeted to have 8,000 to 15,000 cells per well) in 96-well plates to allow accurate quantification of infected cells. Total cell counts per well were enumerated by nuclear stain (Hoechst 33342) and fluorescent virally infected foci were detected 16-24 hours after inoculation with a Cytation 7 Cell Imaging Multi-Mode Reader (BioTek) with Gen5 Image Prime version 3.09. Titers were calculated in GraphPad Prism version 8.4.2 by generating a 4-parameter (4PL) logistical fit of the percent neutralisation at each serial serum dilution. The 50% neutralisation titre (VNT_50_) was reported as the interpolated reciprocal of the dilution yielding a 50% reduction in fluorescent viral foci.

### VSV-SARS-CoV-2 spike variant pseudovirus neutralisation assay

Vesicular stomatitis virus (VSV)-SARS-CoV-2-S pseudoparticle generation and neutralisation assays were performed as previously described^21^. Briefly, human codon optimized SARS-CoV-2 spike (GenBank: MN908947.3) was synthesised (Genscript) and cloned into an expression plasmid. SARS-CoV-2 complete genome sequences were downloaded from GISAID Nucleotide database (https://www.gisaid.org). Sequences were curated and genetic diversity of the spike-encoding gene was assessed across high quality genome sequences using custom pipelines. Amino acid substitutions were cloned into the spike expression plasmid using site-directed mutagenesis. HEK293T cells (ATCC CRL-3216) were seeded (culture medium: DMEM high glucose [Life Technologies] supplemented with 10% heat-inactivated fetal bovine serum [FBS; Life Technologies] and penicillin/ptreptomycin/L-glutamine [Life Technologies]) and transfected the following day with spike expression plasmid using Lipofectamine LTX (Life Technologies) following the manufacturer’s protocol. At 24 hours post-transfection at 37 °C, cells were infected with the VSVΔG:mNeon/VSV-G virus diluted in Opti-MEM (Life Technologies) at a multiplicity of infection of 1. Cells were incubated 1 hour at 37 °C, washed to remove residual input virus and overlaid with infection medium (DMEM high glucose supplemented with 0.7% Low IgG BSA [Sigma], sodium pyruvate [Life Technologies] and 0.5% Gentamicin [Life Technologies]). After 24 hours at 37 °C, the supernatant containing VSV-SARS-CoV-2-S pseudoparticles was collected, centrifuged at 3000xg for 5 minutes to clarify and stored at -80 °C until further use.

For pseudovirus neutralisation assays, Vero cells (ATCC CCL-81) were seeded in 96-well plates in culture medium and allowed to reach approximately 85% confluence before use in the assay (24 hours later). Sera were serially diluted 1:2 in infection medium starting with a 1:40 dilution. VSV-SARS-CoV-2-S pseudoparticles were diluted 1:1 in infection medium for a fluorescent focus unit (ffu) count in the assay of ∼1000. Serum dilutions were mixed 1:1 with pseudoparticles for 30 minutes at room temperature prior to addition to Vero cells and incubation at 37 °C for 24 hours. Supernatants were removed and replaced with PBS (Gibco), and fluorescent foci were quantified using the SpectraMax i3 plate reader with MiniMax imaging cytometer (Molecular Devices). Neutralisation titers were calculated in GraphPad Prism version 8.4.2 by generating a 4-parameter logistical (4PL) fit of the percent neutralisation at each serial serum dilution. The 50% pseudovirus neutralisation titre (pVNT_50_) was reported as the interpolated reciprocal of the dilution yielding a 50% reduction in fluorescent viral foci.

### IFNγ ELISpot

IFNγ ELISpot analysis was performed *ex vivo* (without further *in vitro* culturing for expansion) using PBMCs depleted of CD4^+^ and enriched for CD8^+^ T cells (CD8^+^ effectors), or depleted of CD8^+^ and enriched for CD4^+^ T cells (CD4^+^ effectors). Tests were performed in duplicate and with a positive control (anti-CD3 monoclonal antibody CD3-2 [1:1,000; Mabtech]). Multiscreen filter plates (Merck Millipore) pre-coated with IFNγ−specific antibodies (ELISpotPro kit, Mabtech) were washed with PBS and blocked with X-VIVO 15 medium (Lonza) containing 2% human serum albumin (CSL-Behring) for 1-5 hours. Per well, 3.3 × 10^5^ effector cells were stimulated for 16-20 hours with an overlapping peptide pool representing the vaccine-encoded RBD. Bound IFNγ was visualized using a secondary antibody directly conjugated with alkaline phosphatase followed by incubation with BCIP/NBT substrate (ELISpotPro kit, Mabtech). Plates were scanned using an AID Classic Robot ELISPOT Reader and analysed by ImmunoCapture V6.3 (Cellular Technology Limited) or AID ELISPOT 7.0 software (AID Autoimmun Diagnostika). Spot counts were displayed as mean values of each duplicate. T-cell responses stimulated by peptides were compared to effectors incubated with medium only as a negative control using an in-house ELISpot data analysis tool (EDA), based on two statistical tests (distribution-free resampling) according to Moodie et al.^31,32^, to provide sensitivity while maintaining control over false positives.

To account for varying sample quality reflected in the number of spots in response to anti-CD3 antibody stimulation, a normalisation method was applied to enable direct comparison of spot counts/strength of response between individuals. This dependency was modelled in a log-linear fashion with a Bayesian model including a noise component (unpublished). For a robust normalization, each normalisation was sampled 1000 times from the model and the median taken as normalized spot count value. Likelihood of the model: log *λ*_*E*_ = *α* log *λ*_*P*_ + log *β*_*j*_ + *σε*, where *λ*_*E*_ is the normalized spot count of the sample, *α* is a stable factor (normally distributed) common among all positive controls *λ*_*P*_, *β*_*j*_ a sample *j* specific component (normally distributed) and *σε* is the noise component, of which *σ* is Cauchy distributed and *ε* is Student’s-t distributed. *β*_*j*_ ensures that each sample is treated as a different batch.

### Flow cytometry

Cytokine-producing T cells were identified by intracellular cytokine staining. PBMCs thawed and rested for 4 hours in OpTmizer medium supplemented with 2 µg/mL DNase I (Roche), were restimulated with a peptide pool representing the vaccine-encoded SARS-CoV-2 RBD (2 µg/mL/peptide; JPT Peptide Technologies) in the presence of GolgiPlug (BD) for 18 hours at 37 °C. Controls were treated with DMSO-containing medium. Cells were stained for viability and surface markers in flow buffer (DPBS [Gibco] supplemented with 2% FCS [Biochrom], 2 mM EDTA [Sigma-Aldrich]) for 20 minutes at 4 °C. Afterwards, samples were fixed and permeabilized using the Cytofix/Cytoperm kit according to manufacturer’s instructions (BD Biosciences). Intracellular staining was performed in Perm/Wash buffer for 30 minutes at 4 °C. Samples were acquired on a FACS VERSE instrument (BD Biosciences) and analysed with FlowJo software version 10.5.3 (FlowJo LLC, BD Biosciences). RBD-specific cytokine production was corrected for background by subtraction of values obtained with DMSO-containing medium. Negative values were set to zero. Cytokine production in Figure 4b was calculated by summing up the fractions of all CD4^+^ T cells positive for either IFNγ, IL-2 or IL-4, setting this sum to 100% and calculating the fraction of each specific cytokine-producing subset thereof.

### Cytokine profiling

Human PBMCs were restimulated for 48 hours with SARS-CoV-2 RBD peptide pool (2 µg/mL final concentration per peptide). Stimulation with DMSO-containing medium served as negative controls. Concentrations of TNF, IL-1β, IL-12p70, IL-4 and IL-5 in supernatants were determined using a bead-based, 11-plex T_H_1/T_H_2 human ProcartaPlex immunoassay (Thermo Fisher Scientific) according to the manufacturer’s instructions. Fluorescence was measured with a Bioplex200 system (Bio-Rad) and analysed with ProcartaPlex Analyst 1.0 software (Thermo Fisher Scientific). RBD-specific cytokine production was corrected for background by subtraction of values obtained with DMSO-containing medium. Negative values were set to zero.

---

**Extended Data Figure 1.**
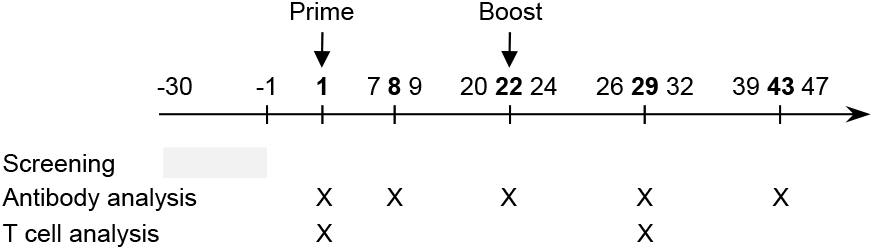
Schedule of vaccination and assessment. Study participants received a prime immunisation with BNT162b1 on Day 1 (all dose levels), and a boost immunisation on Day 22±2 (all dose levels except 60 µg). Serum was obtained on Day 1 (pre-prime), 8±1 (post-prime), 22±2 (pre-boost), 29±3 and 43±4 (post-boost). PBMCs were obtained on Day 1 (pre-prime) and 29±3 (post-boost).

**Extended Data Figure 2.**
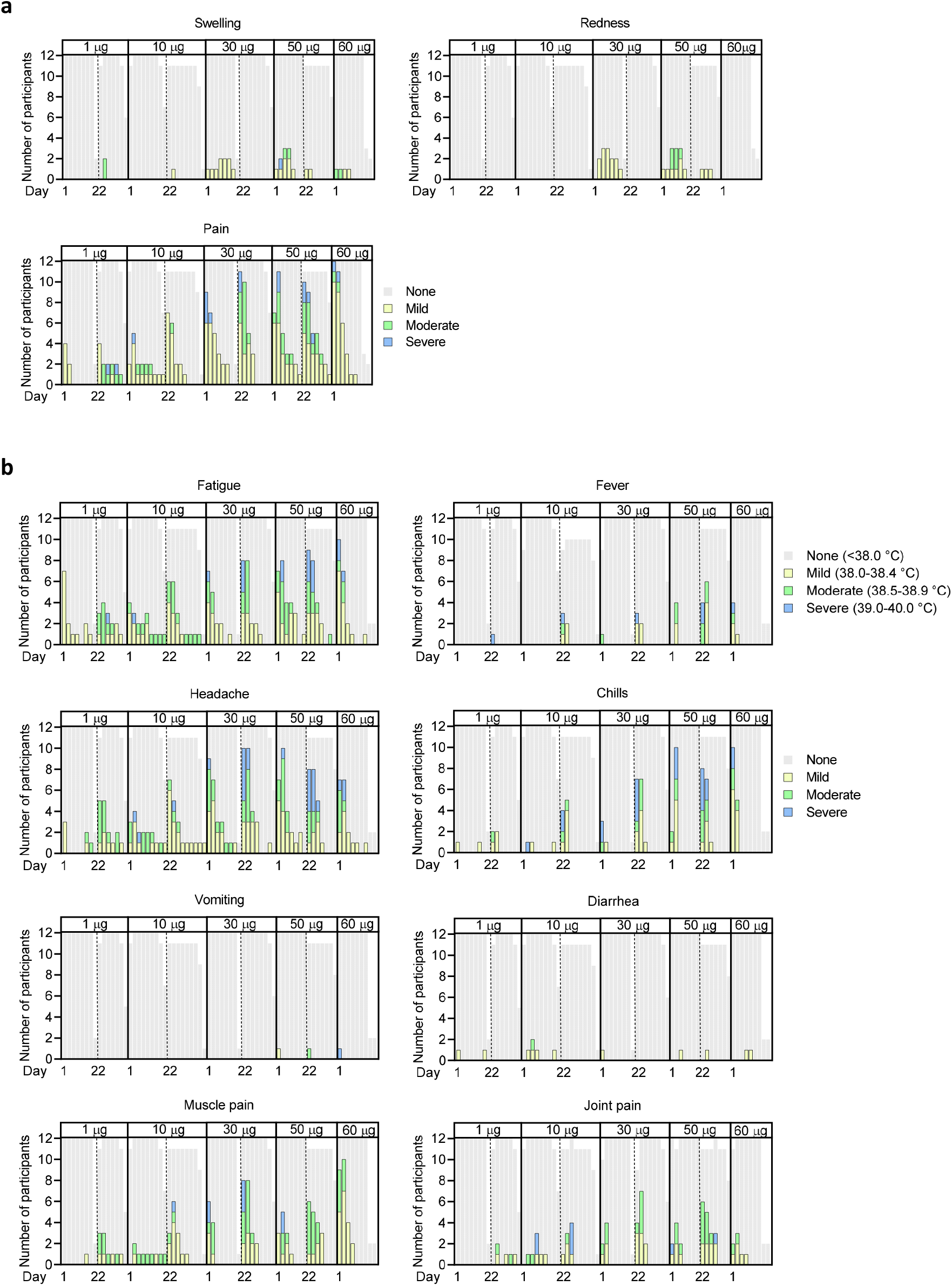
Solicited adverse events. Number of participants with local (**a**) or systemic AEs (**b**). Participants were immunised with BNT162b1 on Days 1 (all dose levels) and 22 (all dose levels except 60 µg) (*n*=12 per group; *n*=11 for 10 µg and 50 µg cohort from Day 22 on; discontinuation of patients due to non-vaccine related reasons; missing data points are indicated). As per protocol AEs were recorded up to 7 days after each immunisation (Days 1-7 and 22-28), and for some participants 1-2 additional days of follow-up were available. Grading of adverse events was performed according to FDA recommendations^33^.

**Extended Data Figure 3.**
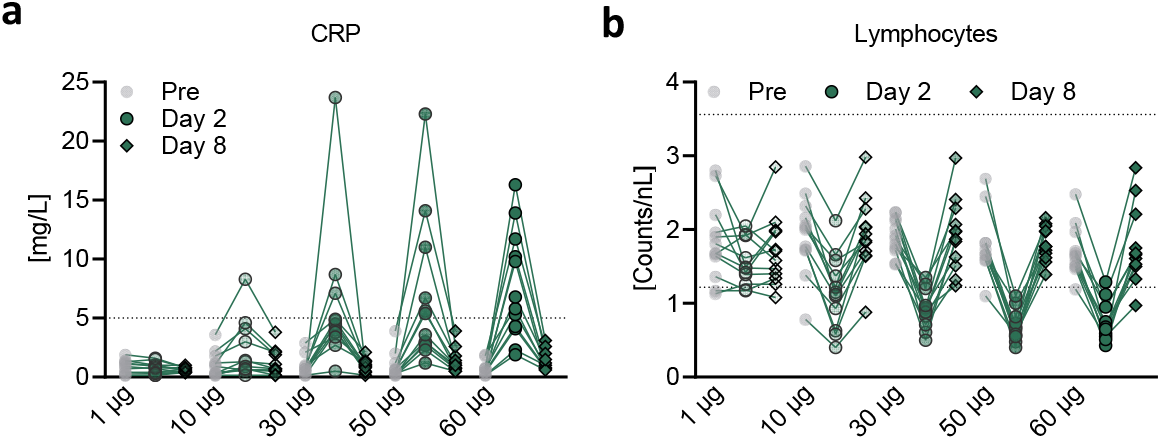
Pharmacodynamic markers. Participants were immunised with BNT162b1 on Days 1 (all dose levels) and 22 (all dose levels except 60 µg). **a**, Kinetics of C-reactive protein (CRP) level and **b**, Kinetics of lymphocyte counts. Dotted lines indicate upper and lower limit of reference range. For values below the lower limit of quantification (LLOQ = 0.3), LLOQ/2 values were plotted (a).

**Extended Data Figure 4.**
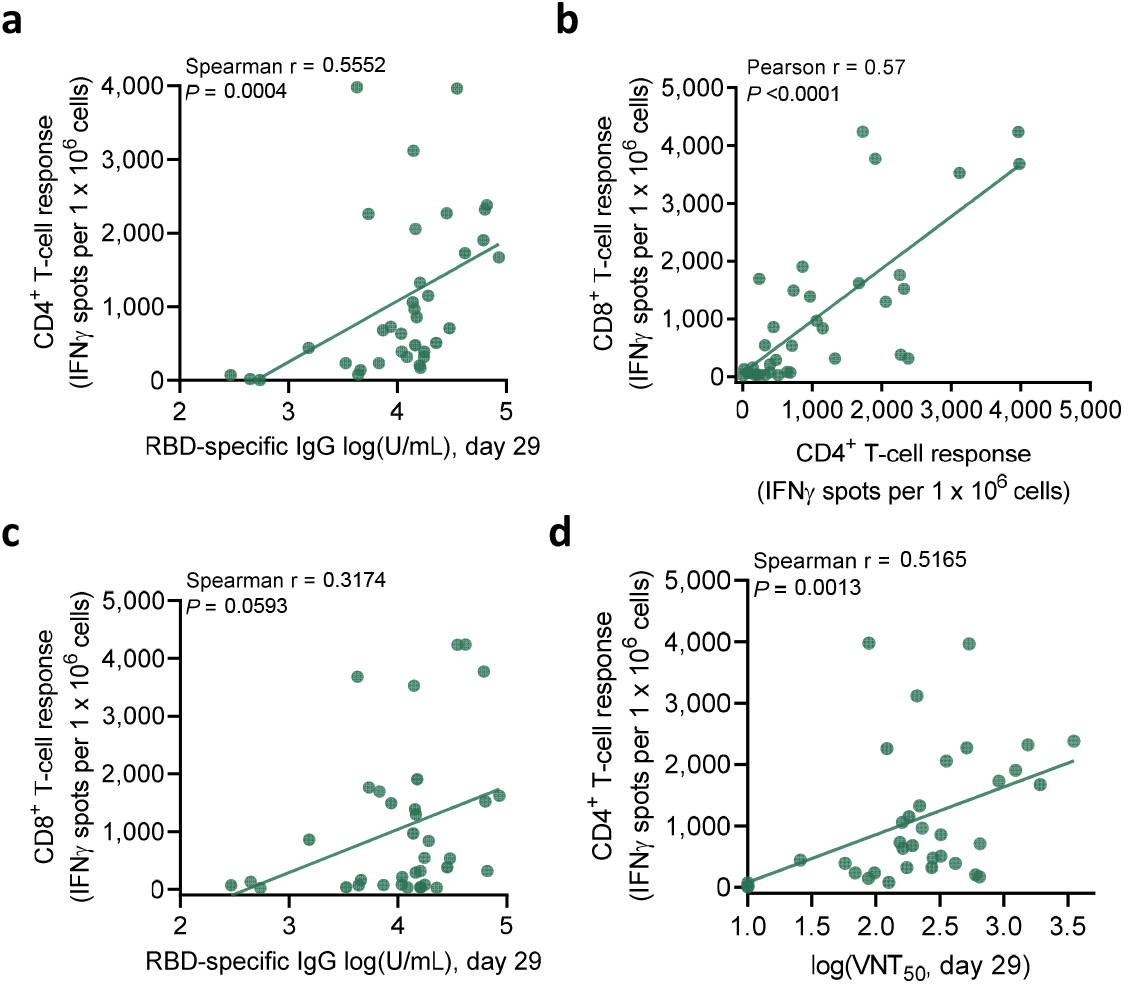
Correlation of antibody and T-cell responses. Participants were immunised with BNT162b1 on Days 1 (all dose levels) and 22 (all dose levels except 60 µg). **a**, Correlation of RBD-specific IgG responses (from Figure 1a) with CD4^+^ T-cell responses on Day 29 (1 and 50 µg, *n*=8 each; 10 and 30 µg, *n*=10 each). Nonparametric Spearman correlation. **b**, Correlation of CD4^+^ with CD8^+^ T-cell responses (as in Figure 3) from Day 29 in dose cohorts 10 to 50 µg. Parametric Pearson correlation. **c**, Correlation of RBD-specific IgG responses (as in Figure 1a) with CD8^+^ T-cell responses (as in Figure 3) on Day 29. Nonparametric Spearman correlation. **d**, Correlation of VNT_50_ (as in Figure 2a) with CD4^+^ T-cell responses (as in Figure 3) in dose cohorts 10 to 50 µg (1 and 50 µg, *n*=8 each; 10 and 30 µg, *n*=10 each).

**Extended Data Table 1.** Subject disposition and analysis sets. Antibody analysis: Values indicate number of participants for whom virus neutralisation assays and RBD binding IgG antibody assays were performed. T-cell analysis: Values indicate number of participants for whom IFNγ ELISpot and flow cytometry (values in parentheses) was performed. N/A: Samples not yet available.

|  | BNT162b1 |  | Antibody analysis |  |  |  |  | T-cell analysis |  |
| --- | --- | --- | --- | --- | --- | --- | --- | --- | --- |
| Cohort | Prime | Boost | Day 1 | Day 8±1 | Day 22±2 | Day 29±3 | Day 43±4 | Day 1 | Day 29±3 |
| 1 µg | 12 | 12 | 12 | 12 | 12 | 12 | 12 | 8 | 8 |
| 10 µg | 12 | 11 | 12 | 12 | 12 | 11 | 11 | 10 (6) | 10 (6) |
| 30 µg | 12 | 12 | 12 | 12 | 12 | 12 | 12 | 10 (7) | 10 (7) |
| 50 µg | 12 | 11 | 12 | 12 | 12 | 11 | 11 | 8 (5) | 8 (5) |
| 60 µg | 12 | N/A | 12 | 12 | 11 | 12 | N/A | N/A | N/A |

